# Prohibit, protect, or adapt? The changing role of volunteers in palliative and hospice care services during the COVID-19 pandemic. A multinational survey (CovPall)

**DOI:** 10.1101/2021.03.28.21254486

**Authors:** Catherine Walshe, Ian Garner, Lesley Dunleavy, Nancy Preston, Andy Bradshaw, Rachel L Cripps, Sabrina Bajwah, Katherine E Sleeman, Mevhibe Hocaoglu, Matthew Maddocks, Fliss EM Murtagh, Adejoke O Oluyase, Lorna K Fraser, Irene J Higginson, on behalf of the CovPall study team

**Affiliations:** International Observatory on End of Life Care, Lancaster University, UK; Wolfson Palliative Care Research Centre, Hull York Medical School, University of Hull, UK; Cicely Saunders Institute of Palliative Care, Policy & Rehabilitation, King’s College, London, UK; The Martin House Research Centre, Department of Health Sciences, University of York, UK

**Keywords:** Palliative care, COVID-19, Volunteers

## Abstract

**Background:** Volunteers are common within palliative care services, and provide support that enhances care quality. The support they provided, and any role changes, during the COVID-19 pandemic are unknown.

**Aims:** To understand volunteer deployment and activities within palliative care services, and to identify what may affect any changes in volunteer service provision, during the COVID-19 pandemic.

**Methods:** Multi-national online survey disseminated via key stakeholders to specialist palliative care services, completed by lead clinicians. Data collected on volunteer roles, deployment, and changes in volunteer engagement. Analysis included descriptive statistics, a multivariable logistic regression, and analysis of free-text comments using a content analysis approach.

**Results:** 458 respondents: 277 UK, 85 rest of Europe, and 95 rest of the world. 68.5% indicated volunteer use pre-COVID-19. These were across a number of roles (from 458): direct patient/family facing support (58.7%), indirect support (e.g. driving) (52.0%), back office (48.5%) and fundraising (45.6%). 11% had volunteers with COVID-19. Of those responding to a question on change in volunteer deployment (328 of 458) most (256/328, 78%) indicated less or much less use of volunteers. Less use of volunteers was associated with being an in-patient hospice, (OR=0.15, 95%CI = 0.07-0.3 p<.001). This reduction in volunteers was felt to protect potentially vulnerable volunteers and with policy changes preventing volunteers from supporting services. However, adapting was also seen where new roles were created, or existing roles pivoted to provide virtual support.

**Discussion and conclusion:** Volunteers were mostly prevented from supporting many forms of palliative care, particularly in in-patient hospices, which may have quality and safety implications given their previously central roles. Volunteer re-deployment plans are needed that take a more considered approach, using volunteers more flexibly to enhance care while ensuring safe working practices. Consideration needs to be given to widening the volunteer base away from those who may be considered to be most vulnerable to COVID-19.

## BACKGROUND

Specialist palliative and hospice care services have proven to be critically important as part of the whole-system management of the COVID-19 pandemic (1–3). They are closely involved in the symptom management of those who are dying or who have challenging symptoms (both dying with COVID-19 and from COVID-19) and, at times, services have provided additional bed capacity to help manage the surge in patient numbers in the wider health care system. Whilst paid staff are central to the provision of palliative and hospice care services, volunteers are also major contributors to the way that high quality, safe services are provided across the world (4). In some services volunteers can be more numerous than paid staff, with one UK survey identifying 1.5 volunteers to every paid member of staff (5), providing a great number of hours of care and support, typically up to 8 hours a week (6). It is estimated that each UK volunteer provides at least £1,500 of value per annum to the organisation (7). Volunteers also offer stability; a Belgian survey identified that most volunteers had been in their current care organisation for at least 6 years (57%), and 36% for over 10 years (6).

Volunteers can support many different aspects of palliative and hospice care across all settings, including in-patient palliative care units, hospital and home palliative care teams, home nursing services and in the community (8, 9). Whilst volunteers traditionally contributed mostly to ‘back office’ functions such as finance or catering, as well as running shops and other fundraising activities, they are increasingly found in patient facing roles (10–13). When providing patient-facing care, typically the focus is on psychosocial support, including existential care, signposting as well as care tasks (5, 6, 14–17). Volunteers complement professional care by being a unique face of care for patients, occupying a liminal space between professionals, family and patients (8, 18, 19).

Care from volunteers is safe, effective, and appreciated by patients (9). Benefits to people who receive care are assumed to include improvements in quality of life and enhancement of wellbeing (9, 11, 15, 19–22), and one study also indicated a survival advantage for those supported by volunteers (23). Volunteers themselves benefit from their volunteering activities reporting that it becomes a major part of their lives (4), changing their own perspectives and values (24–26).

No data are yet available on the impact of the COVID-19 pandemic on volunteers and the role and service they provide to palliative and hospice services during this time. Effective use of volunteers is highlighted as a possible response to the pandemic (27), with calls for mobilising and training a citizen volunteer workforce that is ready and able to connect with patients in need of basic social support (28). It is important that the role of volunteers during the COVID-19 pandemic is understood, given the dependence many palliative care services have on them for quality care provision and to maintain a safe organisation.

## METHODS

### Aim

To understand volunteer deployment and activities within palliative and hospice care services, and to identify what may affect any changes in volunteer service provision, during the COVID-19 pandemic.

### Design

A cross-sectional design, with a single point of data collection using an online multi-national survey of hospice and specialist palliative care providers. This study is reported in accordance with STROBE (29) and CHERRIES (30) statements. This paper is part of the wider CovPall study (1–3) that aims to understand the multi-national specialist palliative care response to COVID-19.

### Population and setting

Service leads were invited to take part in the online survey on behalf of their organisation if they provided a minimum of one of the following specialist palliative care services: in-patient palliative care, hospital palliative care, home palliative care and home nursing.

### Sampling and recruitment

The survey was open to responses from 23/04/2020 to 31/07/2020. Invitations to participate were disseminated through open advertisement and via palliative care and hospice organisations (Sue Ryder, Hospice UK, Scottish Partnership for Palliative Care, Marie Curie, European Association of Palliative Care, Together for Short Lives, and the palliativedrugs.com and www.pos-pal.orgnetwork). Interested eligible services were provided with a link to complete the survey online, together with a participant information sheet. Completion indicated consent.

### Data collection

REDCap (an online web application that allows for the building and managing of surveys and databases) was used to collect data online with closed and free text survey responses, designed to shed light on the context for closed responses. Sites were given the option to enter the data online directly, be emailed the survey to complete and then return electronically, or complete the survey via telephone or video conferencing with a member of the study team. As well as general and COVID-19 related service information (see Appendix 1 for full survey), specific questions were asked about their use of volunteers, and the impact of COVID-19 on volunteers (Table 1).

**Table 1:** Specific survey questions on volunteer use

|  |  |
| --- | --- |
| If you had volunteer roles available within your service, what were they? (tick all that apply) | • Direct patient / family facing support (e.g. befriending, home visits, in-patient unit care, family support groups / visiting etc.)<br>• Indirect patient / family facing support (e.g. reception functions, refreshments, driving /transport etc.)<br>• Back office functions (e.g. finance support, maintenance, gardening etc.)<br>• Fundraising functions (e.g. shop volunteers, lottery etc.) |
|  | Others (a box will open below) |
| Have you had volunteers with suspected or confirmed COVID-19 | Yes/No |
| What impact has this (volunteers with suspected or confirmed COVID-19) on your service? |  |
| Have you changed how your volunteers engage and where? Please give details | Yes/No |
| How would you say you are deploying volunteers compared to before COVID-19? | A lot more<br>Slightly more<br>About the same<br>Slightly less<br>Much less |

### Data analysis

In the quantitative analyses, the primary outcome was a dichotomised variable about volunteer deployment post-pandemic (a lot more/slightly more/about the same *vs* slightly less/much less volunteer use), collapsed from the initial five-point scale for those services that answered this question, hereafter more or less volunteer use. The relationship between these two categories of volunteer use during the pandemic and a number of potential explanatory variables (service funding model; type of service provided; whether adult/child service; number of confirmed or suspected COVID-19 cases; PPE shortages; staff shortages; whether service changes were made; whether services perceived themselves to be busy; and geography (UK/Europe/Rest of World) were explored using frequency counts (for dichotomous variables) or median/interquartile range (for continuous non normally distributed variables). Differences between more or less volunteer use for dichotomous variable were assessed using Chi Square (χ^2^) analysis, with Mann-Whitney U (U) t-tests for non-dichotomous data. Sample size (n) is also provided for Mann-Whitney U t-tests. For the multivariable logistic regression model, the dependent variable was change in frequency of volunteer use (with ‘less volunteer use’ as the reference category), with explanatory variables chosen according to significance (p<.05). For each explanatory variable the reference category was the answer ‘no’ for dichotomous variables, and the lowest denominator for non-dichotomous variables (e.g. ‘much less busy’ for staff busyness), and for the outcome the ‘less volunteer use’ was the reference group. Model fit was assessed using BIC. Analysis was conducted in SPSSv26.

For the analysis of free-text comments, data were extracted for the relevant questions in table 1. As is common with free text data from surveys comments tended to be brief, expanding on answers to closed questions (31, 32). After initial familiarisation, a coding framework was developed and applied to the free text data (CW) using a conventional content analysis technique (33).

### Research ethics and approvals

Ethical approval for this study was obtained from King’s College London Research Ethics Committee (21/04/2020, Reference; LRS19/20-18541). The study was registered on the ISRCTN registry (ISRCTN16561225). Completion of survey indicated the participant had consented to the study.

## RESULTS

A total of 458 responses were received, of which 314 (68.5%) indicated they used volunteers pre-pandemic in any role, and with 328 answering the question about deployment during the pandemic (see table 2 for details).

**Table 2.** Descriptive data on volunteer use pre and during the COVID-19 pandemic

|  | All responses<br>(n=458) | Indicated any past<br>volunteer use<br>(n=314) | Answered question<br>about current<br>volunteer<br>deployment (the<br>same or more/less).<br>(n=328) |
| --- | --- | --- | --- |
| **Geography:** |  |  |  |
| UK | 277 (60.5%) | 187 (59.6%) | 195 (59.5%) |
| Europe | 85 (18.6%) | 59 (18.8%) | 62 (18.9%) |
| Rest of World | 95 (20.7%) | 67 (21.3%) | 71 (21.6%) |
| Missing | 1 (0.2%) | 1 (0.3%) | 0 |
| **Pre-pandemic volunteer roles:** |  |  |  |
| Direct support | 269 (58.7%) | 269 (85.6%) | 246 (75.0%) |
| Indirect support | 238 (51.9%) | 238 (75.7%) | 218 (66.4%) |
| Back office | 222 (48.4%) | 222 (70.7%) | 205 (62.5%) |
| Fundraising | 209 (45.6%) | 209 (66.5%) | 189 (58.1%) |
| Others | 51 (11.1%) | 51 (16.2%) | 49 (14.9%) |
| Missing | 0 | 0 | 0 |
| **Volunteers with COVID-19** |  |  |  |
| Yes | 38 (8.3%) | 36 (11.4%) | 36 (10.9%) |
| No | 369 (80.6%) | 260 (82.8%) | 279 (85.0%) |
| Missing | 51 (11.1%) | 18 (5.8%) | 13 (4.1%) |
| **Have you changed how your volunteers engage?** |  |  |  |
| Yes | 280 (61.1%) | 268 (85.3%) | 258 (78.6%) |
| No | 119 (26.0%) | 34 (10.8%) | 64 (19.8%) |
| Missing | 59 (12.9%) | 12 (3.9%) | 6 (1.6%) |
| **How would you say you are deploying volunteers compared to before COVID-19** |  |  |  |
| A lot more | 12 (2.6%) | 11 (3.3%) | 12 (3.6%) |
| Slightly more | 10 (2.2%) | 9 (2.9%) | 10 (3.0%) |
| About the same | 50 (10.9%) | 23 (7.4%) | 50 (15.4%) |
| Slightly less | 29 (6.3%) | 29 (9.3%) | 29 (8.8%) |
| Much less | 227 (49.6%) | 211 (67.2%) | 227 (69.2%) |
| Missing | 130 (28.4%) | 31 (9.9%) | 0 |

Further analyses only include data from the 328 services who responded to the question about volunteer deployment during the pandemic (Table 3). When comparing the 130 participants who did not provide answers on volunteer deployment compared to those who did, participants who did not answer this question had significantly more PPE shortages (χ^2^ = 6.65, p = 0.01), staff shortages (χ^2^ = 4.63, p = .03), and changes to hospital palliative care advanced team settings in response to COVID-19 (χ^2^ = 4.59, p = 0.03). No further significant differences were found.

**Table 3:**
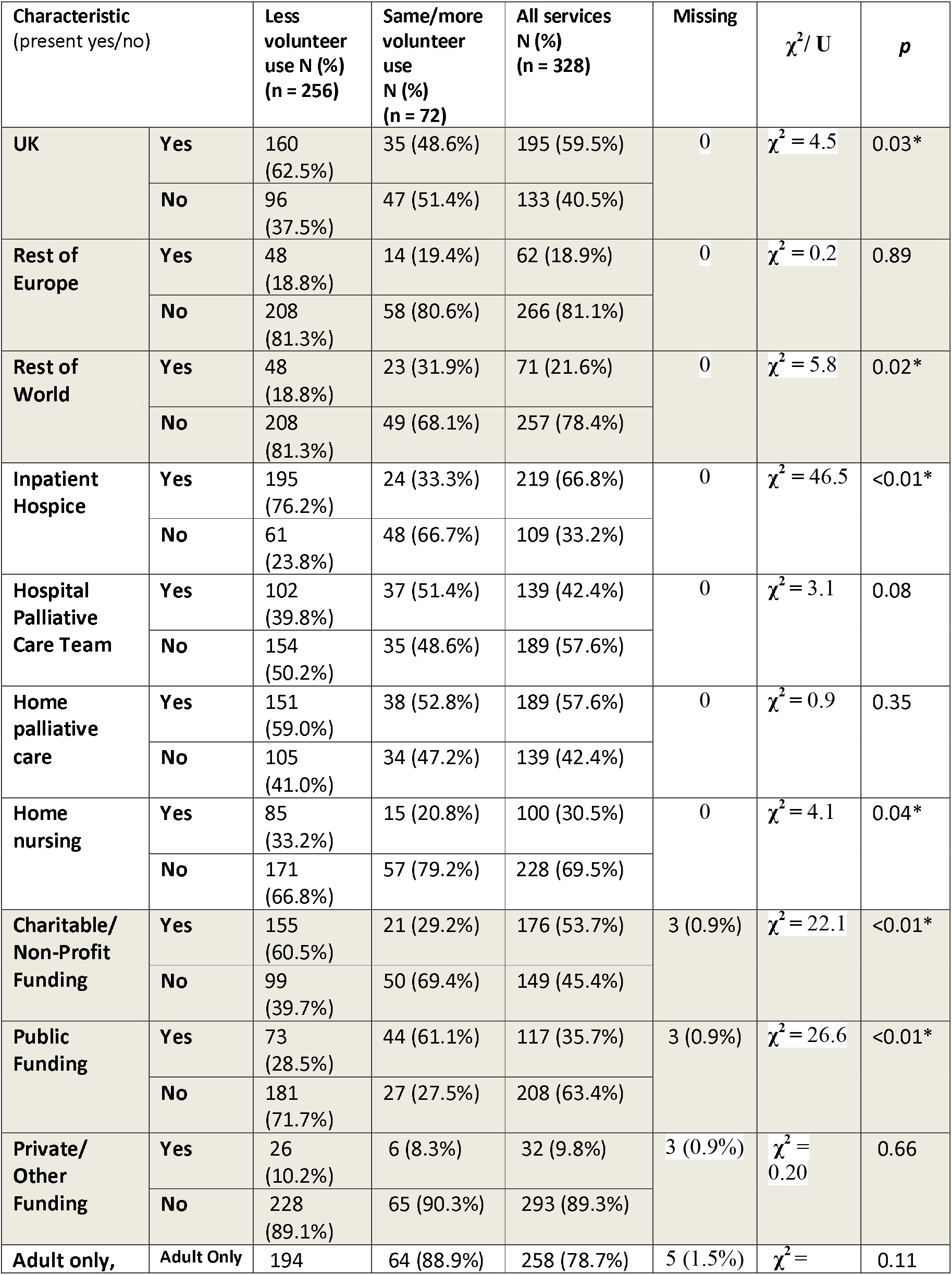
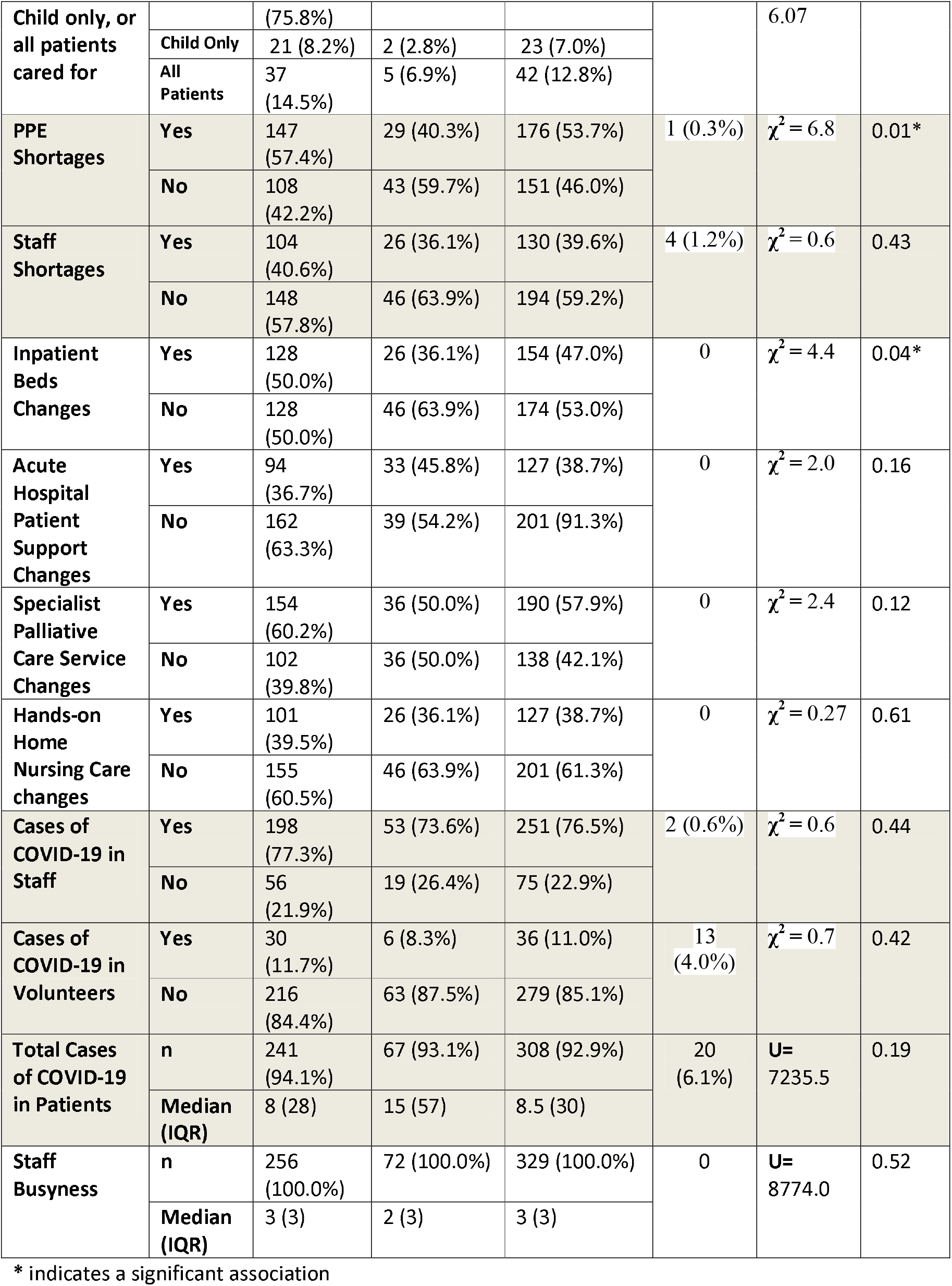
Comparison of characteristics of services indicating more or less volunteer use during the COVID-19 pandemic

The multivariable logistic regression (Table 4) shows that there was a significant association between providing in-patient hospice care and reporting less use of volunteers than usual during the pandemic.

**Table 4:**
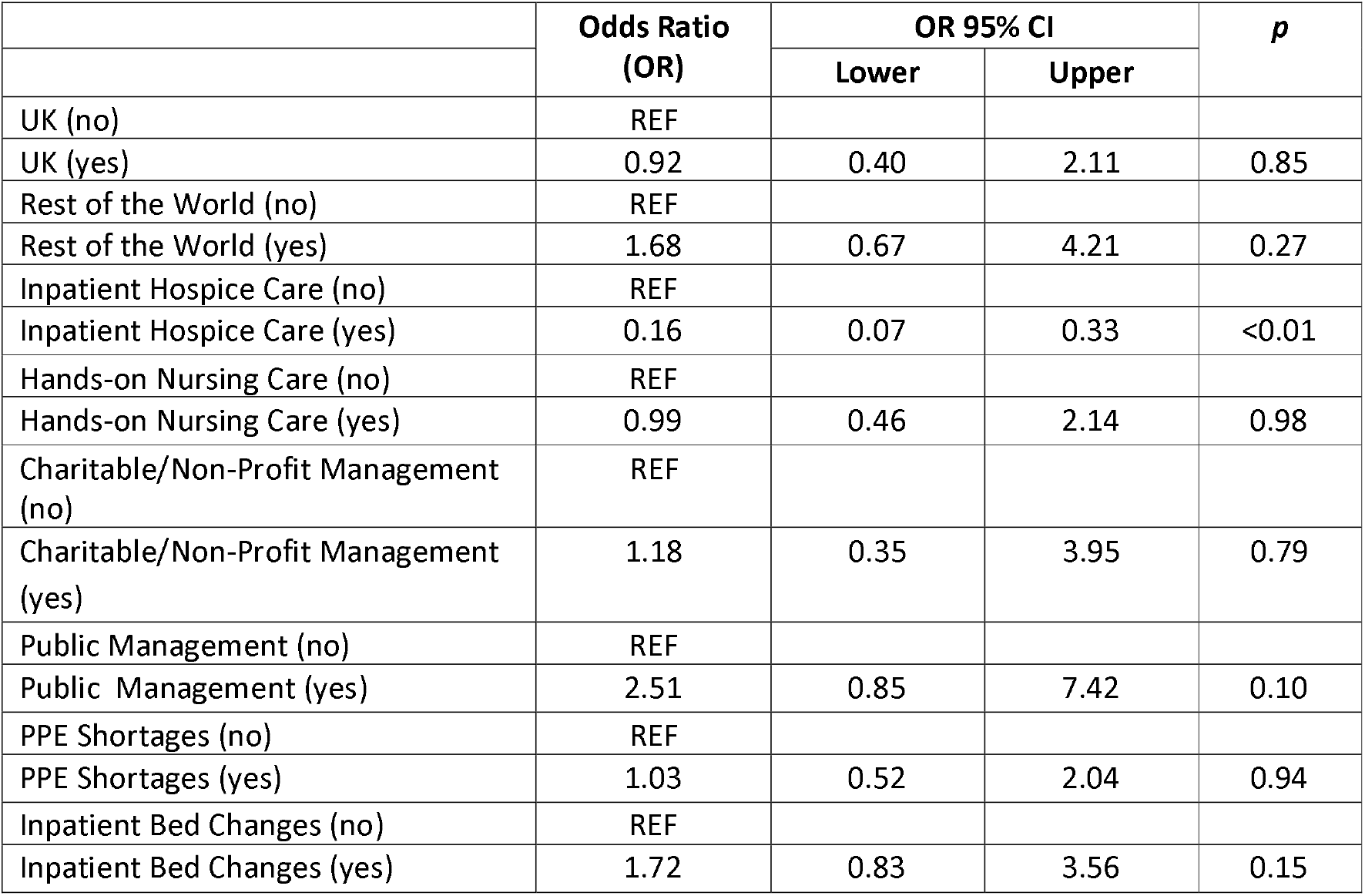
Variables independently associated with less volunteer use during the COVID pandemic

|  | Odds Ratio<br>(OR) | OR 95% CI |  | p |
| --- | --- | --- | --- | --- |
|  |  | Lower | Upper |  |
| UK (no) | REF |  |  |  |
| UK (yes) | 0.92 | 0.40 | 2.11 | 0.85 |
| Rest of the World (no) | REF |  |  |  |
| Rest of the World (yes) | 1.68 | 0.67 | 4.21 | 0.27 |
| Inpatient Hospice Care (no) | REF |  |  |  |
| Inpatient Hospice Care (yes) | 0.16 | 0.07 | 0.33 | <0.01 |
| Hands-on Nursing Care (no) | REF |  |  |  |
| Hands-on Nursing Care (yes) | 0.99 | 0.46 | 2.14 | 0.98 |
| Charitable/Non-Profit Management (no) | REF |  |  |  |
| Charitable/Non-Profit Management (yes) | 1.18 | 0.35 | 3.95 | 0.79 |
| Public Management (no) | REF |  |  |  |
| Public Management (yes) | 2.51 | 0.85 | 7.42 | 0.10 |
| PPE Shortages (no) | REF |  |  |  |
| PPE Shortages (yes) | 1.03 | 0.52 | 2.04 | 0.94 |
| Inpatient Bed Changes (no) | REF |  |  |  |
| Inpatient Bed Changes (yes) | 1.72 | 0.83 | 3.56 | 0.15 |

Services who care for adult patients only were significantly associated with more volunteer use. No other variables were significantly associated with change in volunteer use compared to pre-COVID-19.

Analysis of free-text data identified two overarching themes. First, that of protecting and prohibiting volunteers from contributing in the ways that they did pre-pandemic. Second, that of adaptation, where a minority of services adapted and changed the way they deployed volunteers.

### Protect and prohibit

Our quantitative data demonstrates a large decrease in the use of volunteers. Our free text data illuminates this, identifying that typically volunteers were either prohibited from supporting the service in the way that they usually did, or else because they were protected because they were perceived as particularly vulnerable to the effects of COVID-19. This was both because of local service based policies, or in response to national guidelines about the protection of those who were particularly vulnerable:

> *“Volunteers were temporarily told to stay home across the hospital. Elderly volunteers were told to stay longer periods at home for their protection*.*”* - **Site 478** (*Rest of world, Adult, Hospital*)
>
> *“All volunteer work cancelled due to demographic of majority of volunteers, and concern about exposing them to COVID by charity*.*”* - **Site 99** (*UK, Adult, IPU/Hospital advisory*)
>
> *“No volunteers are called upon to offer their services. This is largely because our volunteers are generally over 65yr and there is fear from their families of undue exposure and risk*.” - **Site 25** (*Rest of world, Adult, IPU*)

Concerns about protecting volunteers from COVID were noted both because of their personal vulnerabilities, the concerns of their families, and of affecting the institution’s reputation if a volunteer contracted COVID-19 as a result of their involvement in the organisation. Institutional policies were often changed to directly prohibit volunteers from enacting their roles:

> *“Early corporate steer - no volunteers in the hospital*.*”* - **Site 188** (*UK, Adult, Hospital*)
>
> *“The hospital/trust have altered their policy on this. No ward volunteers, volunteers redeployed to e*.*g. distributing donations”* - **Site 250** (*UK, Adult, Hospital*)

As well as protecting or prohibiting the volunteers themselves, preserving and prioritising both the distribution of PPE when there were shortages, and also the integrity of the site, was also important, with sites favouring so called ‘essential’ staff as opposed to volunteers. Despite most services reporting that they used volunteers in some capacity pre-pandemic, concerns about supporting and supervising volunteers during the pandemic also contributed to reductions in their deployment, with many of them not considering volunteers to be essential staff:

> *“Reduced ward-based volunteers to preserve PPE and reduce the footfall on the ward*.*”* - **Site 59** (*UK, IPU/Hospice*)
>
> *“Due to changes in services and changed working practices unable to support and supervise volunteers. Only essential staff working in the hospice hence no volunteers attending when families are in*.*”* - **Site 52** (*UK, Children, IPU*)

Such decisions had a knock-on effect on staffing across the organisation, with staff being re-deployed to support functions previously run by volunteers:

> *“Staff have been deployed so duties such as reception are being supported by staff*.*”* **Site 47** (*England, Adult, IPU/Hospital/Home*)

### Adaptation

Some services had identified safe ways of adapting roles, or developed new functions that volunteers could more safely fulfil during the pandemic. This included support, befriending and bereavement roles, often delivered remotely. Other roles included services such as driving, delivering, shopping and gardening. Occasionally completely new roles were identified which could include those directly arising as a result of the pandemic (e.g. making scrubs), but also coordination and information sharing roles. An example is the pivot to telephone or virtual support for patients already known to the organisation, and using skills that volunteers had already developed in existing in-person roles:

> *“We’ve asked all existing befriending or bereavement type volunteers to offer telephone support and soon to offer Facebook group bereavement support. We’ve asked Compassionate Neighbours to offer support to care home residents. We hope to set up a bereavement telephone helpline for any resident in [name of region] (and once lockdown eases we will need more volunteers to help act as a listening ear)*.*”* **Site 56** (*UK Adult, IPU/Hospital/community*)
>
> *“Now utilizing ‘buddy program’ where volunteers can call individuals and do a check in and offer support to help with social isolation and bridge the gap from quarantine at home and the community”* - **Site 373** (*US/Adult/Hospital/community*)

More rarely, services imagined a completely new role for volunteers that hadn’t been fulfilled in-person previously. Examples included both new remote roles, such as facilitating the completion of care plans, or in-person roles such as providing hands-on nursing care:

> *“New volunteers helping patients with myCMC [coordinate my care – a care planning initiative]. Volunteers calling GP practices to get them to complete CMC plans. Volunteers calling care homes to navigate them through the creation of myCMC plans for their residents*.*”* - **Site 76** (*UK, Adult, IP/Hospice*)
>
> *“Additional volunteer training provided early on so that volunteers can provide basic patient care. This has been a really popular move for both volunteers and staff and will continue and develop*.*”* - **Site 187** (*UK, Adult, IPU*)

## DISCUSSION

Palliative and hospice care services that had previously been reliant on a large volunteer body to support care often experienced a large decline in the presence of volunteers during the early phase of the COVID-19 pandemic, primarily due to their active withdrawal or suspension by the organisation to protect volunteers and focus on a core staff team. Some palliative and hospice care organisations instituted new roles for volunteers, or moved existing roles to a remote way of working, but these appeared uncommon. In-patient hospices appeared particularly vulnerable to seeing reductions in volunteer use.

The management of risk within an organisation is important, but challenging to undertake at speed in a pandemic situation when new and previously unknown risks are presenting themselves. COVID-19 has highlighted the vulnerabilities of organisations, and led to challenging dilemmas about how to manage care standards in a crisis (34). It is perhaps understandable in this context that a simple solution to manage the risks associated with volunteers is to rapidly curtail their activities, particularly in small organisations that are high users of volunteers, such as many in-patient hospices. Writing plans and procedures to manage volunteers during a pandemic is possibly not an organisational priority. This has also happened previously, such as the suspension of volunteers during Avian flu (35). However, it must be recognised that in such a volunteer-rich specialty that this also carries risk. There is evidence that responding to COVID-19 has strained the palliative care workforce(36), and surges in demand for end-of-life care have exposed and exacerbated underlying gaps in access to specialty-trained physicians and teams, palliative care medications, and bereavement support for patients and families.(37) At a time like this, not having a plan to use what can be a particularly common, valuable, knowledgeable, and committed resource such as volunteers, potentially adds to, rather than avoids, the risks an organisation faces. Some services, however, did not curtail volunteer activities, but were able to respond more flexibly, and innovate rapidly. It is hard to unpick why they were outliers in this mode of service delivery, given the generally flexible, responsive and innovative nature of their general response to the pandemic(1).

It is likely that a major factor in the rapid cessation or curtailment of the use of volunteers was the perception, or reality, of many volunteers being particularly vulnerable to the effects of COVID-19 because of their age. Concerns were likely to be highlighted because of the large degree of uncertainty surrounding this new disease(38). We know that volunteers are predominantly older people (6). However, it is also argued that this view is potentially discriminatory, and that the capacity of older people must be better used. Whilst assumptions may have been made about the technological capability of older people to switch to a remote form of operation, there is evidence that so called ‘silver surfers’ or ‘digital immigrants’ do use technology and can adapt rapidly to using it in ways that are appropriate to their age group (39, 40). It is critically important that we now work to shape future policies (and training) to optimally engage the resources of our aging population, and not unintentionally discriminate against those who are older as policies and procedures change.(41)

There is evidence that volunteers do not always feel informed about the organisation of patient care, or feel the organisation consistently takes their opinion into account. (42) It is likely that volunteers themselves may have had the ability and capacity to produce the needed plans to enable new ways of working, if engaged and asked, although this may be difficult to do at speed and with competing priorities. Certainly, we know that some have argued for new roles for volunteers during the pandemic such as virtual volunteering (27, 28, 43). Some areas where volunteering is deeply embedded, such as in Kerala, have managed to emphasise community participation as part of their response to COVID, which includes supporting palliative care patients (44). This is not just seen in low-middle income countries, for example the calls for new volunteers in the UK such as the NHS volunteering scheme were responded to by 750, 000 people. Here there is a paradox, volunteers are both seen as central to the response of a community or organisation, but equally not fully integrated into the response of the organisations for which they volunteer, not seen as ‘essential’, and rapidly sidelined due to restrictive policies? For volunteers themselves, it is likely rarely about the tasks themselves, but about volunteering being a fundamental response; a desire to help. Their compassionate response to palliative care needs during COVID should not be put to one side, but ways found of ensuring that they can again become a central and fundamental part of palliative and hospice care provision.

### Strengths and limitations

This was a large, multi-national survey with closed and free-text design giving insight and understanding. The way that this survey was constructed, with single responses covering multiple modes of service provision meant that it was not always possible to fully understand the impact of volunteer changes on specific types of services. The survey was also completed by service leads, and hence reflects their views, not those of volunteers themselves. There were many services that did not provide information on change in volunteer deployment, and they may represent a different type of service. The survey was open for completion over a period of months, and it therefore also represents different times, in different countries, of the experience of the first wave of COVID-19. The temporal sequence of events is not known (e.g. whether an increase in COVID-19 cases triggered a reduction in volunteer use). Free text comments, whilst commonly given, were often short with little context, so it was not always possible to fully interpret justifications for decisions made.

## Conclusions

Volunteers, previously central to the support of many forms of palliative care, were mostly absent from organisations immediate response to COVID-19, particularly in-patient hospices. At a time where staffing has been affected by deployment changes and illness, this lack of a previously stable support may have affected both the quality and safety of care. Flexible deployment plans need to be developed that protect volunteers, whilst still enabling them to have a role supporting care. Consideration needs to be given to widening the volunteer base away from those who may be considered to be most vulnerable to COVID-19.

## Supporting information

Appendix 1 Full survey

Strobe statement

## Data Availability

Applications for use of the survey data can be made for up to 10 years, and will be considered on a case by case basis on receipt of a methodological sound proposal to achieve aims in line with the original protocol. The study protocol is available on request. All requests for data access should be addressed to the Chief Investigator via the details on the CovPall website (https://www.kcl.ac.uk/cicelysaunders/research/evaluating/covpall-study, and) and will be reviewed by the Study Steering Group.

## The CovPall study group

### CovPall Study Team

Professor Irene J Higginson (Chief Investigator), Dr Sabrina Bajwah (Co-I), Dr Matthew Maddocks (Co-I), Professor Fliss Murtagh (Co-I), Professor Nancy Preston (Co-I), Dr Katherine E Sleeman (Co-I), Professor Catherine Walshe (Co-I), Professor Lorna K Fraser (Co-I), Dr Mevhibe B Hocaoglu (Co-I), Dr Adejoke Oluyase (Co-I), Dr Andrew Bradshaw, Lesley Dunleavy, Ian Garner and Rachel L Cripps.

### CovPall Study Partners

Hospice UK, Marie Curie, Sue Ryder, Palliative Outcome Scale Team, European Association of Palliative Care (EAPC), Together for Short Lives and Scottish Partnership for Palliative Care.

## Declarations

### Authorship

IJH is the grant holder and chief investigator; KES, MM, FEM, CW, NP, LKF, SB, MBH and AO are co-applicants for funding. IJH and CW with critical input from all authors wrote the protocol for the CovPall study. MBH, AO, RC and LD co-ordinated data collection and liaised with centres, with input from IJH, FEM, CW, NP and LKF. IG, MH, AO, LF and CW analysed the data. All authors had access to all study data, discussed the interpretation of findings and take responsibility for data integrity and analysis. CW and IF drafted the manuscript. All authors contributed to the analysis plan and provided critical revision of the manuscript for important intellectual content.

### Ethics and consent

Research ethics committee approval for this study was obtained from King’s College London Research Ethics Committee (21/04/2020, Reference; LRS19/20-18541). ISRCTN16561225. Completion of survey indicated the participant had consented to the study.

### Data sharing

Applications for use of the survey data can be made for up to 10 years, and will be considered on a case by case basis on receipt of a methodological sound proposal to achieve aims in line with the original protocol. The study protocol is available on request. All requests for data access should be addressed to the Chief Investigator via the details on the CovPall website (https://www.kcl.ac.uk/cicelysaunders/research/evaluating/covpall-study, and⍰)⍰and will be reviewed by the Study Steering Group.

## Acknowledgements

This study was part of CovPall, a multi-national study, supported by the Medical Research Council, National Institute for Health Research Applied Research Collaboration South London and Cicely Saunders International. We thank all collaborators and advisors. We thank all participants, partners, PPI members and our Study Steering Group. We gratefully acknowledge technical assistance from the Precision Health Informatics Data Lab group (https://phidatalab.org) at National Institute for Health Research (NIHR) Biomedical Research Centre at South London and Maudsley NHS Foundation Trust and King’s College London for the use of *REDCap* for data capture.

