## Appendix 1 Full survey for "Prohibit, protect, or adapt? The changing role of volunteers in palliative and hospice care services during the COVID-19 pandemic. A multinational survey (CovPall)"

### COVPALL COLLABORATION SURVEY

Improving palliative care for people with COVID-19 by sharing learning

Thank you for agreeing to complete this survey. We are trying to find out about how palliative care and hospice services are changing as a result of the COVID-19 pandemic. This is important as the disease is new and hospices/palliative care services are changing how they work and there is an opportunity to learn from each other.

We realise you are very busy right now, and so we have tried to balance collecting the information that patients, policy makers and services think is most helpful, with keeping the questionnaire as short as we can.

The questionnaire has 7 sections, and should take no longer than 30 minutes to complete, although it may depend on how much additional/open comments you wish to share. We will consider everything that you say. Your reply will help us. Although the grouped results will be shared, we will not name your unit unless you ask us to.

You can pause the questionnaire by clicking the "Save & Return Later" button at the bottom of each page. You will be given a code to enable you to continue later. (There should be a "returning" option at the top right of this page.) If you wish to correct errors after you have clicked "Submit" please with CovPall in the subject line.

If you would like help in completing the survey, or would prefer someone to read the questions to you while you give the answers over the telephone / zoom call, please send your contact details to with CovPall in the subject line.

If you have any concerns about this questionnaire or this study please. CovPall is led by Professor Irene Higginson of the Cicely Saunders Institute, with a multiprofessional team of partners from different organisations and backgrounds. Patient representatives have contributed to this questionnaire and our plans. For more information see https://www.kcl.ac.uk/cicelysaunders (specific page later).

This study has been granted ethical approval by the PNM Research Ethics Subcommittee of King's College London, code LRS-19/20-18541.

Please tick all responses that apply.

#### 1. ABOUT YOU AND YOUR SERVICE

1.1 Contact email of the person completing the survey ________________________________
(We need this information so we can help you get back into the survey, or help you to complete it, if you have difficulty)

1.2 Name of the person completing the survey ________________________________

1.3 Date ________________________________
(DD-MM-YYYY)

1.4 Country

◯ England
◯ Scotland
◯ Wales
◯ N Ireland
◯ Australia
◯ Belgium
◯ Canada
◯ Germany
◯ Ireland
◯ Italy
◯ Poland
◯ New Zealand
◯ Other (a box will open)
(A regions option may appear)

1.4a Country (please specify) ______________________________

1.4b English Regions

◯ North East
◯ North West
◯ Yorkshire and The Humber
◯ East Midlands
◯ West Midlands
◯ East
◯ London
◯ South East
◯ South West

1.4c Welsh region

◯ N Wales
◯ W Wales
◯ SE Wales

1.4d Scottish region

◯ Fife, Lothian, Borders, Dumfries & Galloway
◯ Greater Glasgow & Clyde, Ayrshire & Arran, Lanarkshire, Forth Valley
◯ Tayside, Grampian, Western Isles, Highland, Orkney and Shetland

1.4e Region ______________________________

1.5 Your role

◯ Medical director / lead medical clinician
◯ Nurse director / lead nurse clinician
◯ Other (a box will open below)

1.5a Please specify your role ______________________________

---

#### 2. ABOUT THE SERVICES YOU USUALLY OFFERED BEFORE THE COVID-19 PANDEMIC. Please tick all answers that apply unless indicted

2.1 Types of patients cared for

☐ Adults ☐ Children

2.2 In what settings did you provide palliative care services

(additional questions will open for each choice)

☐ In-patient hospice / palliative care unit
☐ Hospital palliative care advisory team
☐ Specialist palliative home care service (supporting / consulting about care for patients at home and/or in the community)
☐ Providing hands on nursing care at home / in the community (e.g. hospice@home, pall@home)
(Tick all that apply)

#### Questions for in-patient hospice / palliative care unit

2.2a Number of beds ________________
(Must be a number)

2.2b Approximate number of new patients seen annually ________________
(Must be a number)

2.2c Normal hours of admitting patients ________________
(E.g. 9:30-17:00)

#### Questions for hospital palliative care advisory team

2.2d Approximate number of new patients seen annually ________________
(Must be a number)

2.2e Did you support
☐ Acute hospitals
☐ Community hospitals

2.2f Normal hours of accepting new referrals ________________
(E.g. 9:30-17:00)

2.2g Did you offer 24/7 support for your patients ☐ Yes

#### Questions for specialist palliative home care service

2.2h Approximate number of new patients seen annually ________________
(Must be a number)

2.2i Did you support patients in care homes ◯ Yes ◯ No

2.2j Normal hours of accepting new referrals ________________
(E.g. 9:30-17:00)

2.2k Did you offer 24/7 support for your patients ☐ Yes

#### Questions for providing hands on nursing care at home / in the community

2.2l Approximate number of new patients seen annually ________________
(Must be a number)

2.2m Normal hours of accepting new referrals ________________
(E.g. 9:30-17:00)

2.2n Did you offer 24/7 support for your patients ☐ Yes

#### Additional information about services offered before the COVID-19 pandemic

2.3 Normal hours of accepting new referrals ________________________

2.4 Did you offer bereavement services ◯ Yes ◯ No

(if yes - additional questions will open)

2.4a Bereavement services / support usually provided and to whom ________________________

2.4b Did you offer bereavement services only to families / friends of patients who had been cared for by your service ◯ Yes ◯ No

2.4c Whom did you offer support to ________________________

2.4d Did you use a risk assessment tool to help you decide how to target bereavement services ◯ Yes ◯ No

2.5 Other services provided, e.g. Day care, rehabilitation, lymphoedema, outpatient clinics - please detail

________________________

2.6 If you had volunteer roles available within your service, what were they

Tick all that apply

- ☐ Direct patient / family facing support (e.g. befriending, home visits, in-patient unit care, family support groups / visiting etc.)
- ☐ Indirect patient / family facing support (e.g. reception functions, refreshments, driving / transport etc.)
- ☐ Back office functions (e.g. finance support, maintenance, gardening etc.)
- ☐ Fundraising functions (e.g. shop volunteers, lottery etc.)
- ☐ Others (a box will open below)

2.6a Please specify the other volunteer roles ________________________

2.7 Did you use remote consultations to help support patients in your care or for education before the COVID-19 Pandemic

Tick all that apply (an example box will appear)

- ☐ Telephone support for education
- ☐ Telephone support for clinical care
- ☐ Telehealth / video support / e-learning for education
- ☐ Telehealth / video support / e-learning for clinical care

2.7a Please give a brief example of the use of remote consultations ________________________

2.8 Is your service managed as a unit that is

- ◯ Charitable / non-profit
- ◯ Public
- ◯ Private
- ◯ Other (a box will open below)

2.8a Please explain your unit type ________________________

2.8b what percentage of your funding was usually from the NHS ________________________
(Must be a number (0-100))

2.9 How well would you say your service was integrated with other NHS primary or secondary care services in your area (e.g. with hospitals, primary care etc.)

Please rank the level of integration: from 0 (no integration at all, no connections) to 10 (very well integrated / close working and planning)

0 5 10

(Place a mark on the scale above)

2.10 Any comments on integration ________________

2.11 Is there anything else you want to tell us about how your service operates that you think is important for us to know ________________

#### 3. EXPERIENCE WITH SUSPECTED OR CONFIRMED CASES OF COVID-19.

**Please tick all that apply**

**Please tell us about people you have encountered with suspected or confirmed COVID-19**

3.1 Have you had any patients with confirmed (by test) cases of COVID-19 ○ Yes ○ No

(additional questions will open if you tick yes)

3.1a Approximately how many (confirmed cases) ________________
(By the date of completing this survey)

3.1b Which services were they in

Tick all that apply

- ☐ In-patient hospice / palliative care unit
- ☐ Home palliative care service
- ☐ Acute hospital
- ☐ Care home
- ☐ Other (a box will open below)

(Tick all that apply)

3.1c Please specify the other services ________________

3.2 Have you had any patients with suspected (untested but with clinical diagnosis/symptoms) of COVID-19 ○ Yes ○ No

(additional questions will open if you tick yes)

3.2a Approximately how many (suspected cases) (by the date of completing this survey) ________________
(Must be a number)

3.2b Which services were they in

- ☐ In-patient hospice / palliative care unit
- ☐ Home palliative care service
- ☐ Acute hospital
- ☐ Care home
- ☐ Other (a box will open below)

(Tick all that apply)

3.2c Please specify the other services ________________

3.2d Of the patients you have seen with suspected or confirmed COVID-19 would you say that they were

☐ Patients who are severely ill or dying due mainly to COVID-19
☐ Patients with pre-existing illnesses / co-morbidities as well as COVID-19 who are severely ill or dying
☐ Patients known to your service already who now have COVID-19
☐
(Tick all that apply)

3.3 Have you had any family members / close friends of your patients who had suspected or confirmed COVID-19

◯ Yes ◯ No

3.4 Have you had staff with suspected or confirmed COVID-19

(if yes - additional questions will open)

◯ Yes ◯ No

3.4a Were the staff

☐ Nurses
☐ Physicians
☐ Allied health professionals, managed
☐ Reception / administrative staff
☐ Managers
☐ Others (a box will open below)
(Tick all that apply)

3.4b Please specify the other staff

__________________________________

3.4c What impact has this had on your service

__________________________________

3.5 Have you had volunteers with suspected or confirmed COVID-19

(if yes - an additional question will open)

◯ Yes ◯ No

3.5a What impact has this had on your service

__________________________________

#### 4. HOW HAVE YOUR SERVICES CHANGED IN RESPONSE TO COVID-19

4.1 Have your services changed

◯ Yes ◯ No

4.2 Would you say overall you are more busy or less busy that before the COVID-19 Pandemic

◯ A lot more busy
◯ Slightly more busy
◯ About the same
◯ Slightly less busy
◯ Much less busy
(Tick the answer that best applies)

4.3 Why is this

__________________________________________

4.4 Have you lost staff from your service who have been moved to help the NHS elsewhere

(if yes - a box for details will open)

◯ Yes ◯ No

4.4a Please give details (lost staff)

__________________________________

4.5 Have you had staff offered to help your service from health services elsewhere ◯ Yes ◯ No

(if yes - a box for details will open)

4.5a Please give details (offered staff) __________________________________

4.6 Have you changed how your staff work ◯ Yes ◯ No

(if yes - a box for details will open)

4.6a Please give details (how work) __________________________________

4.7 Have you changed where your staff work (e.g. home working) ◯ Yes ◯ No

(if yes - a box for details will open)

4.7a Please give details (where work) __________________________________

4.8 Have you changed how your volunteers engage and where ◯ Yes ◯ No

(if yes - a box for details will open)

4.8a Please give details (changed volunteers) __________________________________

#### Use of virtual technologies

4.9 Would you say that you are using virtual technologies (e.g. zoom / teams etc.) with patients and families

◯ A lot more
◯ Slightly more
◯ About the same
◯ Slightly less
◯ Much less
(Tick the answer that best applies)

4.10 Would you say that you are using virtual technologies (e.g. zoom / teams etc.) with colleagues

◯ A lot more
◯ Slightly more
◯ About the same
◯ Slightly less
◯ Much less
(Tick the answer that best applies)

4.11 What have been the difficulties of using virtual technologies __________________________________

4.12 What has worked well when using virtual technologies __________________________________

#### Advance care planning

4.13 Would you say you are now involved directly with patients / families in advance care planning

◯ A lot more
◯ Slightly more
◯ About the same
◯ Slightly less
◯ Much less
(Tick the answer that best applies)

4.14 Would you say you are now involved advising / supporting others and / or educating about advance care planning

◯ A lot more
◯ Slightly more
◯ About the same
◯ Slightly less
◯ Much less
(Tick the answer that best applies)

4.15 In what ways (if any) have you changed how you are supporting advance care planning

__________________________________________

4.16 What would you say are the main challenges for advance care planning during the COVID-19 pandemic

__________________________________________

#### Bereavement support

4.17 Would you say that you provide more or less bereavement support than before

◯ A lot more
◯ Slightly more
◯ About the same
◯ Slightly less
◯ Much less
(Tick the answer that best applies)

4.18 What would you say are the main challenges for bereavement support during the COVID-19 pandemic

__________________________________________

#### Other support

4.19 How are you supporting patients with COVID-19 who are from more disadvantaged sociodemographic communities (e.g. areas with greater poverty, poor housing, homelessness)

__________________________________________

4.20 Have you encountered patients or families with COVID-19 who are from black and minority ethnic groups

◯ Yes ◯ No

(if yes - a box for details of differences will open)

4.20a Are there any differences in how you are supporting or reaching them

__________________________________________

4.21 Are there any groups (e.g. with different religions, cultures) where you have found supporting the individual needs of people affected by COVID-19 is particularly challenging

__________________________________________

#### Effects on patients who do not have COVID-19

4.22 How has COVID-19 changed how you are supporting the types of patients (e.g. with symptoms and progressive illness) that you would usually support __________________________________

4.23 How has COVID-19 changed how you are supporting the families / those important to patients that you would usually support __________________________________

#### 5. CHANGES IN SPECIFIC SERVICES, E.G. IN-PATIENT, HOSPICE, VOLUNTEERS

The next questions ask about some specific changes that might have occurred, please answer only the sections that apply to your services

5.1 Have there been changes in these areas

(additional questions will open for each choice)

- ☐ In-patient beds in your own service
- ☐ How you provide support for patients in acute hospitals
- ☐ How you provide support for patients in their own homes
- ☐ How you provide support for patients in care homes (including nursing homes)

(Tick all that apply)

#### Changes in in-patient beds in your service

5.1a What changes were there in how you used your beds (if any) ______________________________

5.1b Number of beds

- ◯ Increased
- ◯ Stayed about the same
- ◯ Decreased

5.1c Any changes to admission criteria (if so what was the change) ______________________________

5.1d Any changes to out of hours admissions (e.g. evenings / weekends - if so what was the change) ______________________________

#### Changes in how you provide support for patients in acute hospitals

5.1e Numbers of patients needing support

- ◯ Increased
- ◯ Stayed about the same
- ◯ Decreased

5.1f Would you say your face to face contact with patients / their family members is in general

- ◯ A lot more
- ◯ Slightly more
- ◯ About the same
- ◯ Slightly less
- ◯ Much less

5.1g Would you say your face to face contact with staff is

- ◯ A lot more
- ◯ Slightly more
- ◯ About the same
- ◯ Slightly less
- ◯ Much less

5.1h Would you say your telephone / remote connection advice / support is

- ◯ A lot more
- ◯ Slightly more
- ◯ About the same
- ◯ Slightly less
- ◯ Much less

5.1i Have you changed how your team is organized (e.g. supporting patients with and without COVID-19) ______________________________

5.1j Have you changed your working hours (and in what way) ______________________________

5.1k Have you changed your working practices (and in what way) ______________________________

#### Changes in how you provide support for patients in their own homes

5.1l Numbers of patients needing support

- ◯ Increased
- ◯ Stayed about the same
- ◯ Decreased

5.1m Would you say your face to face contact with patients / their family members is

- ◯ A lot more
- ◯ Slightly more
- ◯ About the same
- ◯ Slightly less
- ◯ Much less

5.1n Would you say your face to face contact with staff is

- ◯ A lot more
- ◯ Slightly more
- ◯ About the same
- ◯ Slightly less
- ◯ Much less

5.1o Would you say your telephone / remote connection advice / support is

- ◯ A lot more
- ◯ Slightly more
- ◯ About the same
- ◯ Slightly less
- ◯ Much less

5.1p Have you changed how your team is organized (e.g. supporting patients with and without COVID-19) ______________________________

5.1q Have you changed your working hours (and in what way) ______________________________

5.1r Have you changed your working practices (and in what way) ______________________________

5.1s Have you changed how medicines are given in the community (e.g. who sets up syringe drivers / families administering medicines)

◯ Yes ◯ No

(if yes - a box for details will open)

5.1t Please give details (changed how medicines are given in the community) ______________________________

#### Changes in how you provide support for patients in care homes (including nursing homes)

5.1u Numbers of patients needing support
- ◯ Increased
- ◯ Stayed about the same
- ◯ Decreased

5.1v Would you say your face to face contact with patients / their family members is
- ◯ A lot more
- ◯ Slightly more
- ◯ About the same
- ◯ Slightly less
- ◯ Much less

5.1w Would you say your face to face contact with staff is
- ◯ A lot more
- ◯ Slightly more
- ◯ About the same
- ◯ Slightly less
- ◯ Much less

5.1x Would you say your telephone / remote connection advice / support is
- ◯ A lot more
- ◯ Slightly more
- ◯ About the same
- ◯ Slightly less
- ◯ Much less

5.1y Have you changed how your team is organized (e.g. supporting patients with and without COVID-19) ________________________________

5.1z Have you changed your working hours (and in what way) ________________________________

5.1α Have you changed your working practices (and in what way) ________________________________

#### Changes to how you are supporting families / those important to patients

5.2 How would you say you are supporting families / those important to patients compared to before
- ◯ A lot more
- ◯ Slightly more
- ◯ About the same
- ◯ Slightly less
- ◯ Much less

5.3 Have you changed how you contact and work with families / those important to patients
◯ Yes ◯ No
(If yes, a box for details will open)

5.3a Please give details (changed how contact and work with families)
________________________________________

#### Changes to how you are deploying volunteers

5.4 How would you say you are deploying volunteers compared to before
- ◯ A lot more
- ◯ Slightly more
- ◯ About the same
- ◯ Slightly less
- ◯ Much less

5.5 Have you changed how you deploy volunteers ◯ Yes ◯ No

(if yes - a box for details will open)

5.5a Please give details (changed how contact and work with families) __________________________________

#### 6. CHALLENGES AND INNOVATIONS IN RESPONSE TO COVID-19

We want to know more about the challenges you have faced, their impacts on your services and care, how you have responded to them and what you found to be your successful innovations

Please tick the challenges that you have faced in your service during COVID-19 Pandemic during the past 1 month

6.1 Have you had problems accessing personal protective equipment ◯ Yes ◯ No

(if yes - additional questions will open)

6.1a Please specify what you had a shortage of __________________________________

6.1b What did you do about this __________________________________

6.1c Has this been a problem in the last 7 days
◯ Yes
◯ Sometimes
◯ No

6.2 Have you had a shortage of key medicines ◯ Yes ◯ No

(if yes - additional questions will open)

6.2a Please specify what you had a shortage of __________________________________

6.2b What did you do about this __________________________________

6.2c Has this been a problem in the last 7 days
◯ Yes
◯ Sometimes
◯ No

6.3 Have you had a shortage of other equipment (e.g. syringe drivers) ◯ Yes ◯ No

(if yes - additional questions will open)

6.3a Please specify what you had a shortage of __________________________________

6.3b What did you do about this __________________________________

6.3c Has this been a problem in the last 7 days

◯ Yes
◯ Sometimes
◯ No

6.4 Have you had a shortage of staff ◯ Yes ◯ No

(if yes - additional questions will open)

6.4a Please specify what you had a shortage of

__________________________________

6.4b What did you do about this

__________________________________

6.4c Has this been a problem in the last 7 days

◯ Yes
◯ Sometimes
◯ No

6.5 Have there been other effects on yourself and/or on staff that you think we should know about

__________________________________

6.6 Please tell us about any other challenges and whether or how you overcame them

__________________________________

6.7 What do you foresee will be the biggest challenges for COVID-19 in your service over the next 1-2 months

__________________________________

6.8 What would help you most to overcome these

__________________________________

---

**Now please tell us about your innovations. We are keen to learn what has worked best for you**

6.9 Please tell us about the change in practice or innovation that you think has been most successful to your working

__________________________________

6.10 Why is this

__________________________________

6.11 What would you say were the most important things that made this possible

__________________________________

6.12 Please list any other important changes / innovations you have made

__________________________________

#### 7. SYMPTOM MANAGEMENT

We are interested to learn more about how you are managing symptoms and psychological / emotional problems in our patients and the trajectories of care

#### How long are patients with COVID-19 under your palliative care service

7.1 What was the shortest time in hours

__________________________
(Give number of hours)

7.2 What was the longest time in days

__________________________
(Give number of days)

#### How are you managing symptoms

7.3 Please indicate which of these settings your management refers to (choose only one about which you have most experience)

○ Inpatient hospital ward
○ Inpatient hospital intensive care
○ Community hospital
○ Inpatient hospice / palliative care ward
○ Community

#### The next questions ask about the treatments you are using

7.4 Do you have protocols for symptom management for COVID-19 patients

○ Yes
○ No
○ Unsure
(Yes will ask for details)

7.4a What sources of information did you use

☐ Locally developed guidance
☐ NICE
☐ NHS
☐ Other (a box will open below)

7.4b Please specify the other protocols

__________________________

You can also email us your usual recommendations of any guidance you provide by email to: , marking the email CovPall in the subject line. We are still interested to know how well you find these treatments are working

#### Breathlessness

7.5 Which medicines and therapies do you usually prescribe

__________________________

7.6 How effective do you find these e.g. time to give relief and how well it works

__________________________

#### Agitation

7.7 Which medicines and therapies do you usually prescribe

__________________________________

7.8 How effective do you find these e.g. time to give relief and how well it works

__________________________________

#### Fever / Shivering

7.9 Which medicines and therapies do you usually prescribe

__________________________________

7.10 How effective do you find these e.g. time to give relief and how well it works

__________________________________

#### Cough

7.11 Which medicines and therapies do you usually prescribe

__________________________________

7.12 How effective do you find these e.g. time to give relief and how well it works

__________________________________

#### Pain

7.13 Which medicines and therapies do you usually prescribe

__________________________________

7.14 How effective do you find these e.g. time to give relief and how well it works

__________________________________

#### Other symptoms

7.15 Please provide details of the treatments you are using for any other symptoms you are seeing commonly in COVID-19, especially if this differs from usual palliative care practice

__________________________________

#### Additional comments

7.16 Please provide any additional comments you would like us to be aware of

__________________________________

7.17 Please tell us if you would like us to help you by providing anything else, or any key questions that you think are important to answer

__________________________________

#### Finally

7.18 Please indicate if you would like / are willing to be contacted regarding any of the following (Tick all that apply)

☐ To receive copy of the early reports and our newsletters as the findings emerge
☐ For us to check any information with you
☐ To be acknowledged as responding to this questionnaire (listed along with other services) in the reports and any publications
☐ To participate in a subsequent survey similar to this one in 6-8 weeks time when you may have made more changes or had more experiences
☐ To collect pseudoanonymized data about a small series (around 10) of patients with COVID-19 in your service. This would involve collecting information on symptom severity, on first assessment in palliative care, in around 2 subsequent time points and at discharge or death, to understand more about the symptoms patients experience and their the effective treatments. This would not be a clinical trial, simply recording your practice and views

7.19 If you wish us to use a different Name or Email for the above (instead of the ones already given) please specify here

__________________________________

7.20 How you would like your service acknowledged in any reports, if applicable

____________________________

You will be free to opt out of receiving the updates at any time, your details will not be passed onto other organisations or used for anything other than with your explicit consent above. Your individual responses will remain confidential, they will be analysed pseudonymously by the research team, with your service identified only by a code number unless you explicitly ask us to do otherwise.

#### Thank you for your help at this difficult time
